# Early Evidence and Predictors of Mental Distress of Adults One Month in the COVID-19 Epidemic in Brazil

**DOI:** 10.1101/2020.04.18.20070896

**Authors:** Stephen Xu Zhang, Yifei Wang, Asghar Afshar Jahanshahi, Jizhen Li, Valentina Gomes Haensel Schmitt

## Abstract

**Objective:** We aim to provide early evidence of mental distress and its associated predictors among adults one month into the COVID-19 crisis in Brazil.

**Methods:** We conducted an online survey of 638 adults in Brazil on March 25–28, 2020, about one month (32 days) cross-sectionally after the first COVID-19 case in South America was confirmed in São Paulo. The 638 adults were in 25 states out of the 26 Brazilian states, with the only exception being Roraima, the least populated state in the Amazon. Of all the participating adults, 24%, 20%, and 18% of them were located in Rio de Janeiro state, Santa Catarina state, and São Paulo state respectively.

**Results:** In Brazil, 52% (332) of the sampled adults experienced mild or moderate distress, and 18.8% (120) suffered severe distress. Adults who were female, younger, more educated, and exercised less reported higher levels of distress. Each individual’s distance from the Brazilian epicenter of São Paulo interacted with age and workplace attendance to predict the level of distress. The “typhoon eye effect” was stronger for people who were older or attended their workplace less. The most vulnerable adults were those who were far from the epicenter and did not go to their workplace in the week before the survey.

**Conclusion:** Identifying the predictors of distress enables mental health services to better target finding and helping the more mentally vulnerable adults during the ongoing COVID-19 crisis.

## INTRODUCTION

The first case of COVID-19 in South America appeared in São Paulo in Brazil on February 26, 2020. While the initial cases were imported from Italy to São Paulo – the economic engine of Brazil with a metropolitan population of 22 million, COVID-19 quickly spread across Brazil, reaching 2,433 cases in a month. As cases spread, so did the distress associated with the virus.^1-3^ Research is starting to identify the potential breakout of large-scale mental health issues.^4^ Early pieces of evidence from China and Iran revealed the prevalence of mental health issues among adults during the COVID-19 outbreak.^5-7^

Despite the early evidence from China, countries vary in their medical systems and resources, cultures, the COVID-19 situation, and their restrictive measures,^4^ and hence research can identify the predictors of mental health in individual countries to enable effective identification of mentally vulnerable groups during the COVID-19 crisis.^6,8^ Such evidence in Latin America, especially Brazil, the largest Latin American country with the biggest amount of COVID infections, in remains in its infancy.^9^

### Mental health during the COVID-19 pandemic in Brazil and Latin America

Brazil and Latin America by and large Latin America, have had “some of the highest COVID-19 death rates in the world”.^9^ The proportion of the informal labor market in the region is 54% of all work across Latin America”, yet many of them struggle to maintain their living and had to risk from their social distancing measures to continue with their lives.^9^ Brazil in particular suffered from the malmanagement of the COVID crisis by its President Jair Bolsonaro. In such a context, researchers have found adults in Brazil had elevated levels of anxiety and depression during pandemic,^10,11^ and the high prevalence of mental disorders might be due to the social isolation.^11^ Health care workers reported symptoms of burnout, anxiety, distress and depression.^12^ However, perceived risk of contagion was not a significant predictor of distress.^13^ Being females,^10,11,14,15^ lower socioeconomic status, lower educational levels,^11^ exposure to COVID-19 information, ^11,16,17^ and reduced income during the pandemic were factors related to worse mental health.^17^

In Latin America, A study of 712 healthcare workers in Bolivia, Ecuador, and Peru revealed that older and less educated healthcare workers were less likely to experience anxiety during the COVID-19 crisis.^18^ A survey of 303 Peruvian healthcare workers during the COVID-19 crisis showed that those who were geographically closer to the epicenter of COVID-19 in Peru (Lima) experienced more anxiety and mental distress.^19^ A survey of 252 Ecuadorian health care workers showed that, 82 (32.5%) of them experienced psychological distress, and 71 (28.2%) of them had anxiety disorder.^20^

### Aim

This paper aims to provide early evidence of mental distress and its predictors among adults in Brazil during the onset of the COVID-19 crisis. Building from early research evidence on mental health in China and Iran, where the COVID-19 outbreak occurred earlier,^5,6,21^ we explore several predictors of distress during the COVID-19 crisis in Brazil. In particular, we examine individuals’ distance from São Paulo – the city most affected by COVID-19 in Brazil based on the model of typhoon eye effect. As the COVID-19 crisis continues to impact Brazil, we hope this research identifies useful predictors to help mental health professionals to be more targeted in locating the more mentally vulnerable individuals in the COVID-19 outbreak to provide timely assistance online or via telephone.

## METHODS

### Contexts

The first confirmed case of COVID-19 in South America was a Brazilian who returned from Italy to São Paulo on February 21, 2020.^22^ São Paulo is the biggest city and the economic center of Brazil. Due to its centrality in the Brazilian transportation network, São Paulo also became a center for the spread of COVID-19 in Brazil.^23^ São Paulo had the highest number of confirmed cases in Brazil and was the first city in Brazil to implement a lockdown in an attempt to slow down the spread of the virus on March 22.

### Study design and participants

About one month after the first COVID-19 case in Brazil, we conducted an online cross-sectional survey on March 25–28, 2020 to gather evidence of adults’ distress during the onset of the COVID-19 pandemic in Brazil. During the survey dates, the total confirmed cases in Brazil increased from 2,433 to 3,904, and deaths increased from 57 to 114. On March 25, São Paulo accounted for more than a quarter of the total confirmed cases in Brazil, and this proportion increased to one third on March 28. We developed the survey in English and had it translated into Portuguese with the help of multiple experts who were bilingual in English and Portuguese. Before launching the Portuguese version of the survey, we pre-tested it with five adults from Brazil (not included in the main sample) to revise to reduce any ambiguous words or sentences. The survey was approved by the ethics committee at Tsinghua University (#20200304). The survey was voluntary with the consent from the participants, and we promised the participants confidentiality and anonymity of their responses. In the cover letter of the survey, we provided the brief information about the objective of the study, the procedure, risks and benefits of participations and sought the consent. The online survey has been distributed among 2550 individuals in 50 social groups on Facebook, WhatsApp and Instagram. A total of 638 adults from various parts of Brazil completed our survey (the response rate is 25%). We had participants from all but one of the 26 states in Brazil – Roraima located in the Amazon is the least populated state of Brazil without participating adults. Our of the participants from the 25 Brazilian states, 24% of participants were in Rio de Janeiro state, 20.3% were in Santa Catarina, 18% of them belonged to São Paulo state, 12.9% in Rio Grande do Sul, followed by Minas Gerais at 5.8%, Ceará about 4.9%, and Bahia about 4.7%.

### Measures

We assessed the participants’ socio-demographic characteristics, including gender, age, educational level, the number of children under 18 years old, geographic location, whether they were COVID-19 positive, their exercise hours per day during the past week, and their workplace attendance. Using the participants’ location, we calculated their individual distance from São Paulo, the epicenter of COVID-19 in Brazil, and their distance from the epicenter ranged from 0 to 3,318 km.

We assessed distress using the COVID-19 Peritraumatic Distress Index (CPDI),^24^ which was specifically designed to capture distress during the COVID-19 outbreak. CPDI consists of 24 questions, with the possible score ranging from 4 to 100 (normal: 4–27, mild or moderate: 28–51, severe: 52–100). We had the survey back-translated from English to Portuguese. The Portuguese version of the survey can be found in the online appendix. The CPDI had a Cronbach’s alpha of 0.87 in the Brazil sample.

## RESULTS

### Descriptive findings

Table 1 presents the descriptive findings of the sampled adults. Of the sample, 57.7% (368) were female, 78.7% (502) reported negative for COVID-19, 0.9% (6) reported positive, and 20.4% (130) were unsure whether they had COVID-19. In terms of exercise during the past week, 57.7% of the participants had not exercised; 21.9%, 6.9% and 5.2% of the participants reported exercising 1, 2 and 3 hours per day during the past week respectively; and 4.1% reported exercising more than 5 hours per day.

**Table 1.**
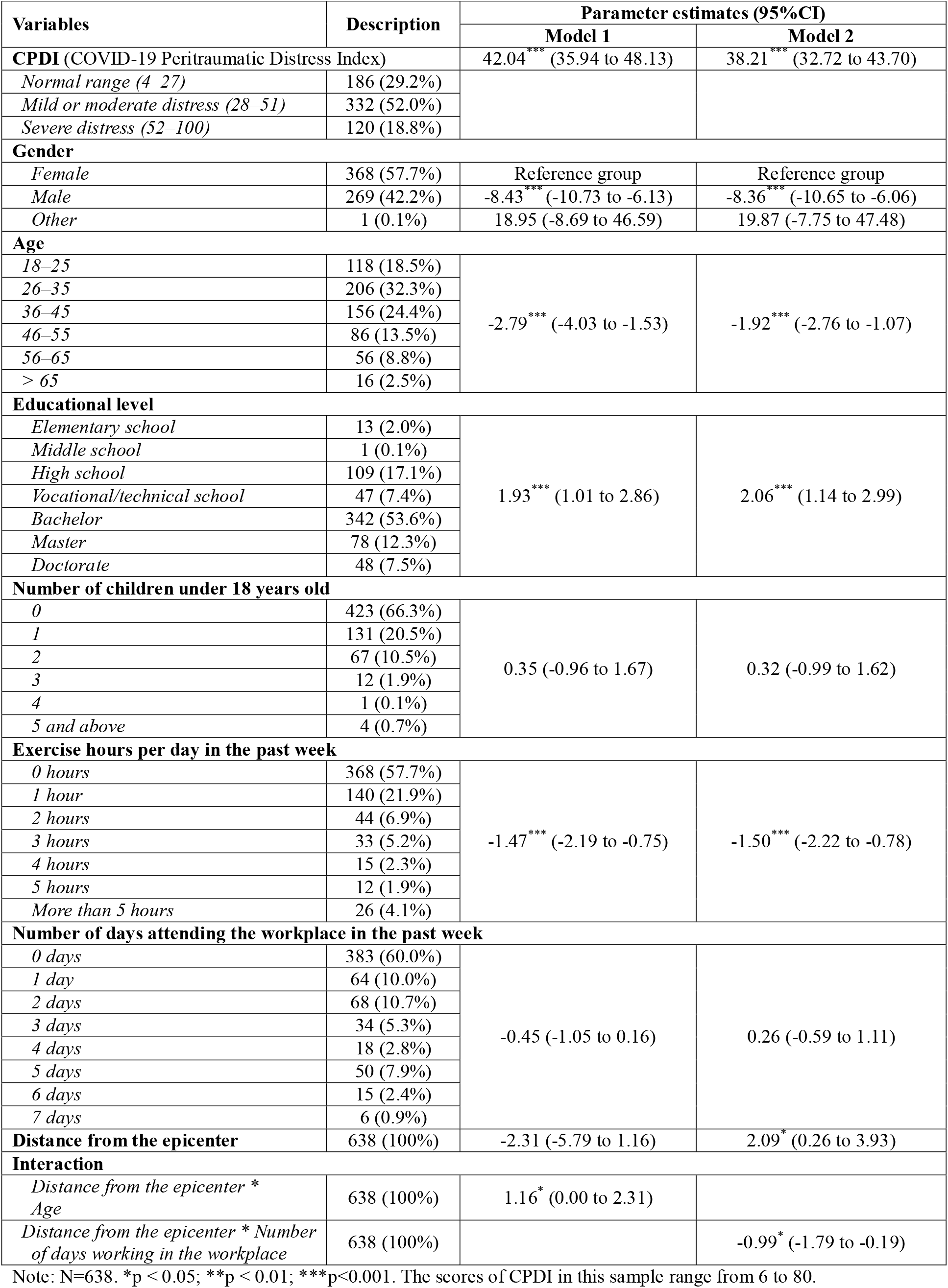
Descriptive findings and predictors of COVID-19 Peritraumatic Distress Index (CPDI)

| Variables | Description | Parameter estimates (95%CI) |  |
| --- | --- | --- | --- |
|  |  | Model 1 | Model 2 |
| CPDI (COVID-19 Peritraumatic Distress Index) |  | 42.04<sup>***</sup> (35.94 to 48.13) | 38.21<sup>***</sup> (32.72 to 43.70) |
| Normal range (4–27) | 186 (29.2%) |  |  |
| Mild or moderate distress (28–51) | 332 (52.0%) |  |  |
| Severe distress (52–100) | 120 (18.8%) |  |  |
| Gender |  |  |  |
| Female | 368 (57.7%) | Reference group | Reference group |
| Male | 269 (42.2%) | -8.43<sup>***</sup> (-10.73 to -6.13) | -8.36<sup>***</sup> (-10.65 to -6.06) |
| Other | 1 (0.1%) | 18.95 (-8.69 to 46.59) | 19.87 (-7.75 to 47.48) |
| Age |  |  |  |
| 18–25 | 118 (18.5%) | -2.79<sup>***</sup> (-4.03 to -1.53) | -1.92<sup>***</sup> (-2.76 to -1.07) |
| 26–35 | 206 (32.3%) |  |  |
| 36–45 | 156 (24.4%) |  |  |
| 46–55 | 86 (13.5%) |  |  |
| 56–65 | 56 (8.8%) |  |  |
| > 65 | 16 (2.5%) |  |  |
| Educational level |  |  |  |
| Elementary school | 13 (2.0%) | 1.93<sup>***</sup> (1.01 to 2.86) | 2.06<sup>***</sup> (1.14 to 2.99) |
| Middle school | 1 (0.1%) |  |  |
| High school | 109 (17.1%) |  |  |
| Vocational/technical school | 47 (7.4%) |  |  |
| Bachelor | 342 (53.6%) |  |  |
| Master | 78 (12.3%) |  |  |
| Doctorate | 48 (7.5%) |  |  |
| Number of children under 18 years old |  |  |  |
| 0 | 423 (66.3%) | 0.35 (-0.96 to 1.67) | 0.32 (-0.99 to 1.62) |
| 1 | 131 (20.5%) |  |  |
| 2 | 67 (10.5%) |  |  |
| 3 | 12 (1.9%) |  |  |
| 4 | 1 (0.1%) |  |  |
| 5 and above | 4 (0.7%) |  |  |
| Exercise hours per day in the past week |  |  |  |
| 0 hours | 368 (57.7%) | -1.47<sup>***</sup> (-2.19 to -0.75) | -1.50<sup>***</sup> (-2.22 to -0.78) |
| 1 hour | 140 (21.9%) |  |  |
| 2 hours | 44 (6.9%) |  |  |
| 3 hours | 33 (5.2%) |  |  |
| 4 hours | 15 (2.3%) |  |  |
| 5 hours | 12 (1.9%) |  |  |
| More than 5 hours | 26 (4.1%) |  |  |
| Number of days attending the workplace in the past week |  |  |  |
| 0 days | 383 (60.0%) | -0.45 (-1.05 to 0.16) | 0.26 (-0.59 to 1.11) |
| 1 day | 64 (10.0%) |  |  |
| 2 days | 68 (10.7%) |  |  |
| 3 days | 34 (5.3%) |  |  |
| 4 days | 18 (2.8%) |  |  |
| 5 days | 50 (7.9%) |  |  |
| 6 days | 15 (2.4%) |  |  |
| 7 days | 6 (0.9%) |  |  |
| Distance from the epicenter | 638 (100%) | -2.31 (-5.79 to 1.16) | 2.09<sup>*</sup> (0.26 to 3.93) |
| Interaction |  |  |  |
| Distance from the epicenter * Age | 638 (100%) | 1.16<sup>*</sup> (0.00 to 2.31) |  |
| Distance from the epicenter * Number of days working in the workplace | 638 (100%) |  | -0.99<sup>*</sup> (-1.79 to -0.19) |

Note: N=638. \*p < 0.05; \*\*p < 0.01; \*\*\*p < 0.001. The scores of CPDI in this sample range from 6 to 80.

The participants reported their workplace attendance by answering the question “how many days did you actually go to work in your office in the past week?”. Of the sample, 60.0% (383) of participants were not in the office at all in the past week, while 28.8% (184) were in the office for fewer than five days last week, 7.9% (50) went to the office for five days, and the remaining 3.3% (21) went for six or seven days.

The mean (SD) score of CPDI in the sample was 37.64 (15.22), higher than the CPDI of 23.65 (15.45) reported in China from January 31 to February 10, 2020.^16^ The difference in the mean values between the samples in Brazil and China is 14.33 (t=23.1; p<0.001; 95%CI: 12.8 to 15.2). The mean CPDI of sampled adults in Brazil is also significantly higher than the mean CPDI of 34.54 (14.92) of adults in Iran on February 28–30, 2020 (t=4.1; p<0.001; 95%CI: 1.6 to 4.6).^6^ Based on the cut-off values of distress in CPDI, 52.0% of sampled adults in Brazil experienced mild or moderate distress, and 18.8% experienced severe distress, compared to 47.0% and 14.1% in Iran and 29.3% and 5.1% in China respectively.

### Predictors of individuals’ COVID-19 Peritraumatic Distress Index (CPDI)

Females experienced more distress than males (β=-8.43, p<0.001, 95%CI: -10.73 to -6.13). Even though COVID-19 has a higher fatality rate in the elderly, younger people reported a higher level of distress (β=-2.79, p<0.001, 95%CI: -4.03 to -1.53). Adults who were more educated (β=1.93, p<0.001, 95%CI: 1.01 to 2.86) and exercised less (β=-1.47, p<0.001, 95%CI: -2.19 to - 0.75) reported a higher level of distress. Family size (p=0.16) and workplace attendance (p=0.63) failed to predict CPDI directly.

We analyzed the relationship between individuals’ distance from the epicenter and CPDI, as well as how this relationship was contingent on their age and the number of days in their workplace during the past week. The relationship between individuals’ distance from the epicenter and their distress depended on individuals’ age (Model 1 of Table 1). First, in Brazil we do observe a “typhoon eye effect” – mental health issues increase with distance from the epicenter, akin to a typhoon, where the effect is stronger in the periphery than in the center. This typhoon eye effect was stronger for older adults (β=1.16, p=0.049, 95%CI: 0.00 to 2.31). We further broke down the typhoon eye effect by adults’ age brackets. The relationship between the distance from the epicenter and distress was significantly positive among older adults (e.g. 46–55 years old: β=2.33, p=0.03, 95%CI: 0.19 to 4.46; 56–65 years old: β=3.49, p=0.03, 95%CI: 0.43 to 6.54; above 65 years old: β=4.65, p=0.03, 95%CI: 0.55 to 8.75).

The relationship between the distance from the epicenter and distress was also contingent on the number of days that the adults went to their workplace during the past week. The number of days in the workplace attenuated the typhoon eye effect in terms of distress (β=-0.99, p=0.02, 95%CI: -1.79 to -0.19), as shown in Model 2 of Table 1. This relationship was significantly positive for adults who did not go to their workplace at all (β=2.09, p=0.03, 95%CI: 0.26 to 3.93), showing the typhoon eye effect. However, this relationship was not significant for adults who went to their workplace for one to five days last week. In particular, the typhoon eye effect (distress increases over distance) turned into the ripple effect (distress decreases over distance) for those who went to their workplace every single day in the last week (β=-4.83, p=0.049, 95%CI: -9.65 to -0.01).

### Predicted scores of individuals’ COVID-19 Peritraumatic Distress Index (CPDI)

Figure 1(a) shows the predicted scores of CPDI by gender, age, education, family size, workplace attendance, and distance from the epicenter. The 95% confidence intervals of CPDI in many groups based on these predictors were higher than the cutoff value of moderate distress at 28. For instance, adults who were female (mean=41.2, 95%CI: 39.7 to 42.6), aged 18–25 (mean=40.9, 95%CI: 39.1 to 42.7), highly educated (individuals with a doctorate degree, mean=42.1, 95%CI: 39.8 to 44.5), and exercised little (for those who did not exercise: mean=39.1, 95%CI: 37.8 to 40.3) all had moderate distress.

**Figure 1.**
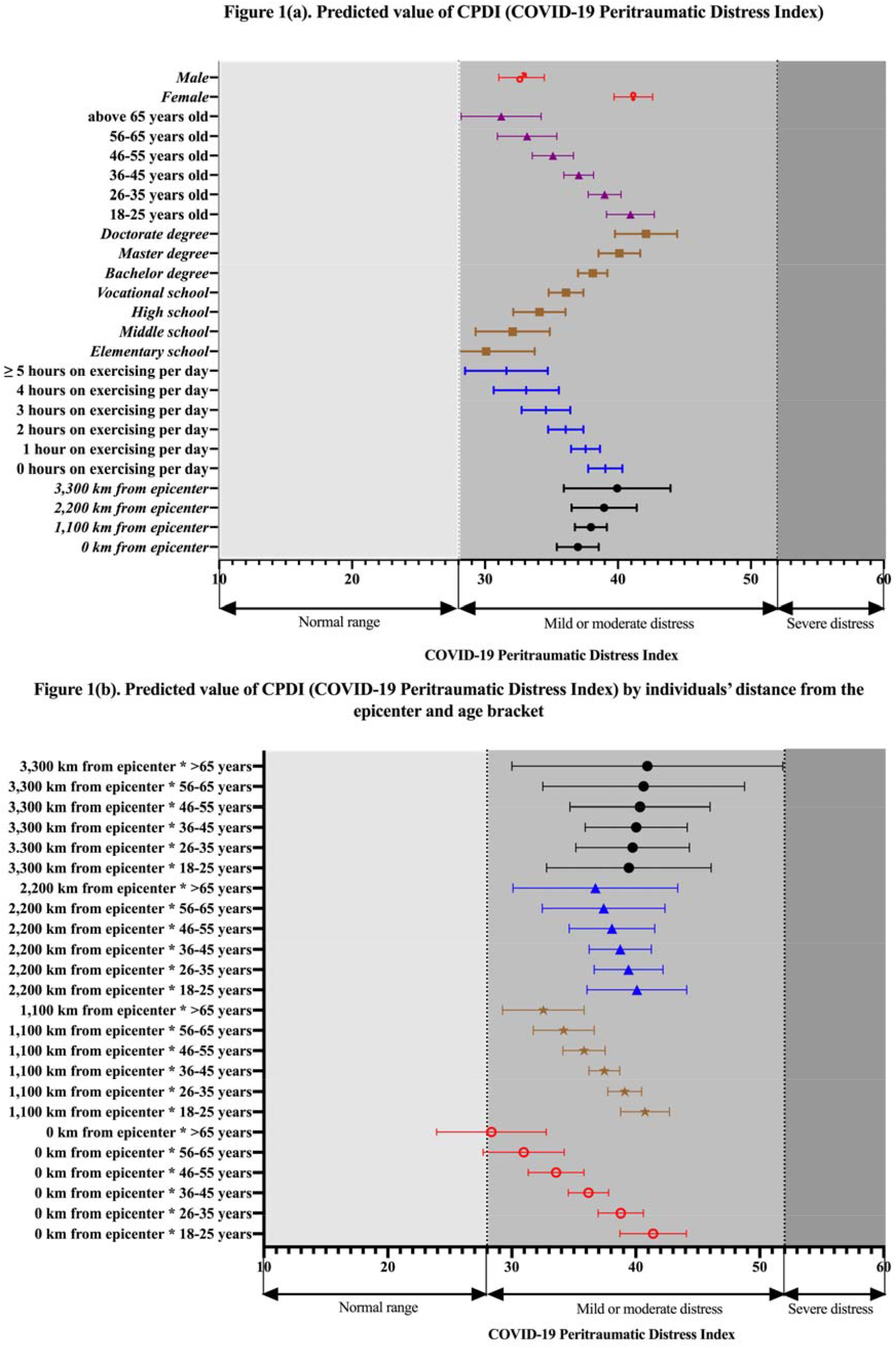
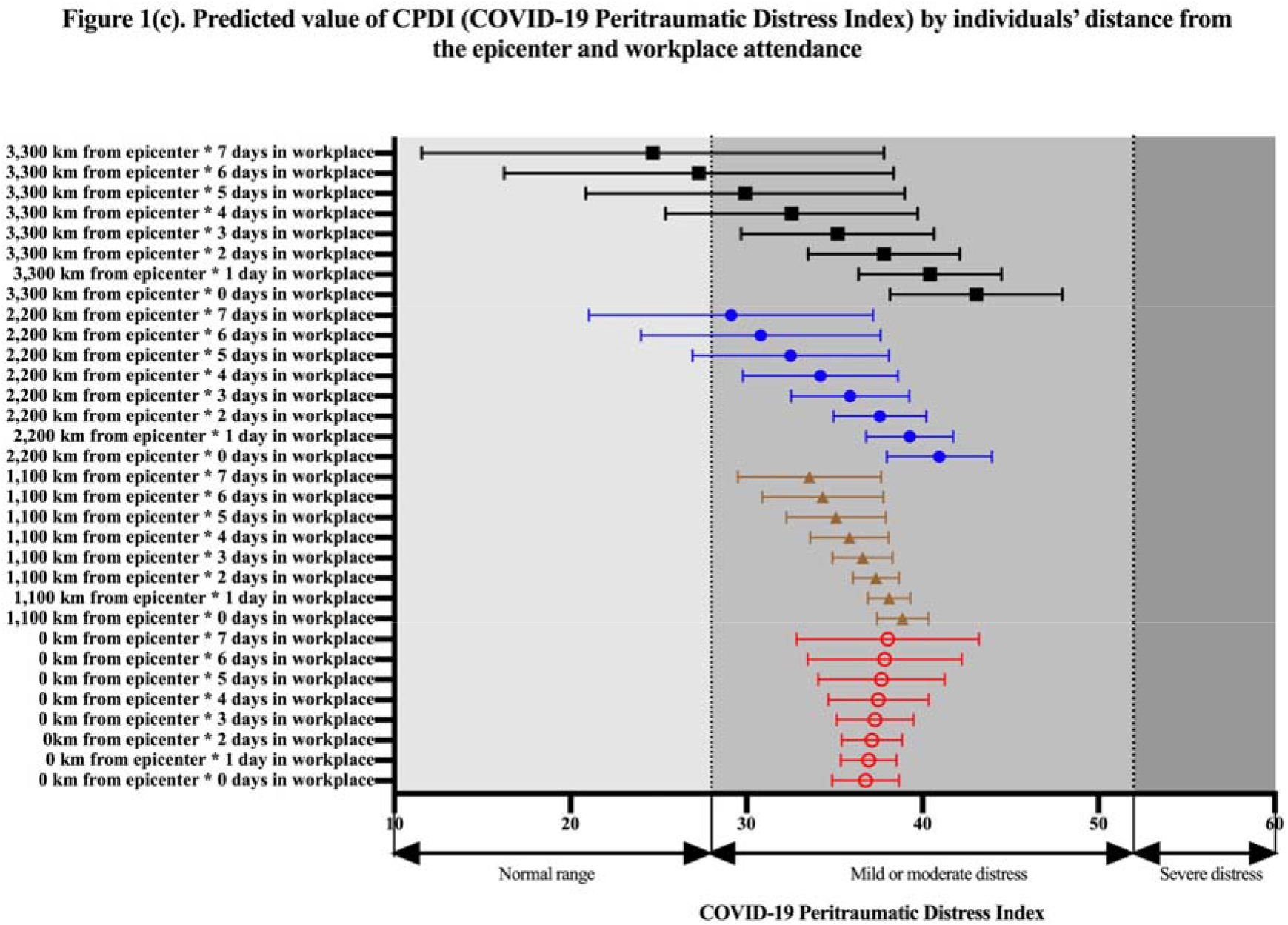
(a) Predicted value of CPDI (COVID-19 Peritraumatic Distress Index) (b) Predicted value of CPDI by individuals’ distance from the epicenter and age bracket (c). Predicted value of CPDI by individuals’ distance from the epicenter and workplace attendance

Since individuals’ distance from the epicenter interacted with their age to predict CPDI level, we plotted the CPDI level based on the interaction of these two factors in Figure 1(b). Individuals aged 18–25 years and who were in the epicenter reported the highest level of distress (mean=41.4, 95%CI: 38.7 to 44.1), and those who were above 65 years old and were 3,300 km from the epicenter in Brazil reported the second highest level of distress (mean=40.9, 95%CI: 30.0 to 51.9). The least distressed group were people older than 65 in the epicenter (mean=28.4, 95%CI: 23.9 to 32.8).

Similarly, Figure 1(c) shows the CPDI level based on the interaction between individuals’ distance from the epicenter and their workplace attendance. The most vulnerable groups during the COVID-19 outbreak were those who were far from the epicenter and did not go to their workplace during the past week (e.g. at 3,300 km from the epicenter: mean=43.1, 95%CI: 38.2 to 48.0; at 2,200 km from the epicenter: mean=41.0, 95%CI: 38.0 to 44.0). The distress level was the lowest among people who lived 3,300 km from the epicenter and attended their workplace every day during the past week (mean=24.7, 95%CI: 11.6 to 37.8).

## DISCUSSION

Our findings reveal a high prevalence of distress among adults during the early stage of the COVID-19 crisis in Brazil. Over half (52.0%) of the adults experienced moderate psychological distress and 18% experienced severe distress. The mean of CPDI of adults in Brazil was also worse than the means in China and Iran. Individuals who were female, younger, more educated, or exercised less had more distress. It is worth noting that two predictors of distress in Brazil, age and education, did not predict distress in the samples in Iran.

The distance from the epicenter is emerging as an interesting predictor of mental health in the crisis literature, and this study found the distance effect depended on individuals’ age and workplace attendance. The positive association between the distance from the epicenter and distress, i.e. the “typhoon eye effect”, was significant only in age groups of 46 years and above. This result might be because the mortality of COVID-19 varies by age group. The typhoon eye effect was significant only among participants who did not attend their workplace. Surprisingly, the effect reversed to become a ripple effect for those who attended their workplace every single day in the last week. There are possible explanations from many perspectives, including the meaning and fulfillment associated with work, more potential social interactions from going out to work, and less time and dependence on information from online and social media. In terms of future research, this study finds that the predictors of distress and their effect during the Covid-19 pandemic In Brazil differ from that in other countries, such as Peru, China, Iran, and Pakistan,^6,19,25,26^ suggesting we need to identify useful predictors of mental health in individual countries during the Covid-19 pandemic, because countries “vary in their medical systems, the availability of personal protective equipment (PPE), cultures, labor and employment conditions, the policies of lockdown, the ease of working from home and maintaining a living in a pandemic, and the information in both mainstream and social media, to name just a few”.^6^

The key contributions of this research are to help identify the predictors of those who are more vulnerable mentally during the COVID-19 crisis to enable more targeted mental health services. We found gender, age, education, exercise, and distance from the epicenter all predicted distress in adults in Brazil during the COVID-19 crisis. In particular, this study shows the predictive effect of the distance from the epicenter varied depending on the age and workplace attendance of each individual. The findings that age and workplace attendance attenuated, and even reversed, the typhoon eye effect is particularly noteworthy to the literature and mental health service providers.

There are several limitations in this study. First, our sampling is not nationally representative, because our aim was to provide rapid evidence on mental health and its predictors to enable rapid screening of the mentally vulnerable in the ongoing COVID-19 outbreak in Brazil. It is worth investigating if the level and the predictors of mental health change as the outbreak continues. Our study aims to provide evidence on the prevalence of distress in the early stage of the Covid-19 crisis, and yet future research may capture how the prevalence and the predictors of mental health may vary over time. Second, Brazil is a large country, and we sampled individuals from 0 to over 3,000 km from São Paulo to cover various regions in Brazil. It remains to be seen to what extent distance from the epicenter is a factor in other countries, most of which are smaller and have their own distinct geographical features.^21^ Third, while the study examines in particular a novel predictor, the distance to the epicenter of the epidemic, as a predictor of the distress experienced by individual adults, we are limited in examining the other predictors. For instance, future research can explore individuals’ work situations, employment sectors (e.g. public, private), self-employment, illegitimate work tasks, seniority at work, whether their workplace downsized, income level, the number of hours they work outside of the home, their health conditions, perceived COVID-19 test availability, belief in COVID as a conspiracy theory,^20, 27, 28^ in the context of Brazil and Latin American countries.

In conclusion, this study provides the early empirical evidence of mental distress and its predictors in adults in Brazil during the COVID-19 crisis. We hope this research not only helps mental health professionals but also encourages more research on mental health conditions and predictors during the COVID-19 crisis in Brazil, Latin America, and beyond.^29, 30^

## Data Availability

Available after publications in a journal

## Acknowledgment

We acknowledge the support of National Natural Science Foundation of China [grant number 71772103]

## The Covid-19 Peritraumatic Distress Index (CPDI) in Portugalese

Selecione a frequência das atividades abaixo na última semana

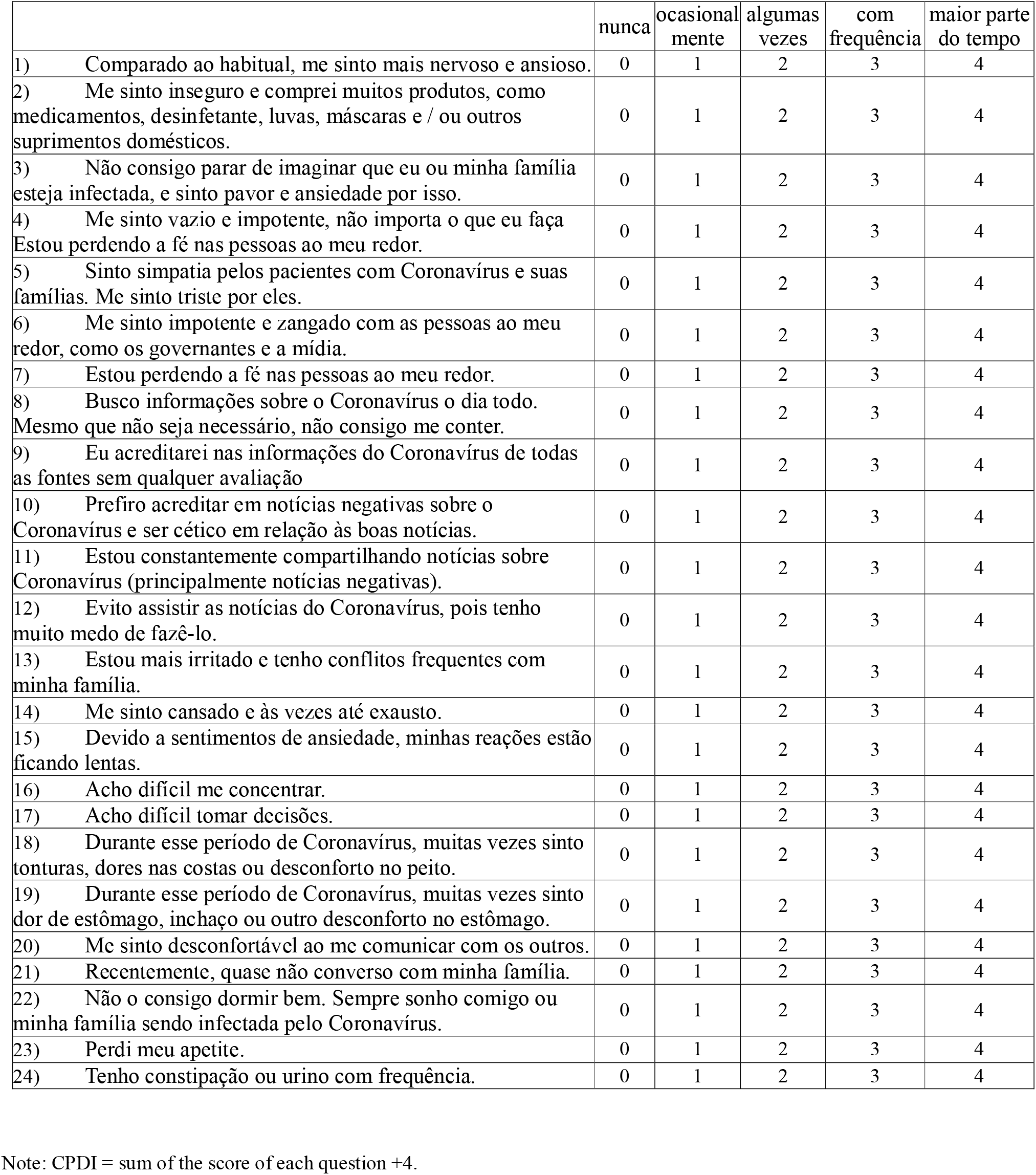

